# Evaluating Clinical Note Deidentification Tools and Transformer Transferability between Public and Private Data from the US Department of Veterans Affairs

**DOI:** 10.1101/2025.03.21.25323520

**Authors:** Hannah Eyre, Qiwei Gan, Mengke Hu, Annie Bowles, Johnathan Stanley, Jianlin Shi, Scott L. DuVall, Patrick R. Alba

**Author notes:** **Corresponding Author:** Patrick Alba.

## Abstract

1

**Background:** Deidentification of clinical notes is critical for enabling secondary use of electronic health records (EHRs) while maintaining patient privacy. Existing deidentification tools vary in methodology, with rule-based, hybrid, and transformer-based approaches offering different trade-offs in performance. However, most evaluations of these tools are conducted on publicly available datasets, limiting their generalizability to private healthcare systems such as the U.S. Department of Veterans Affairs (VA).

**Methods:** We introduce an annotated corpus of 1,000 VA clinical notes and evaluated the performance of four open-source deidentification tools (Physionet Deid, Philter, CliniDeID, and Stanford Deid) using precision, recall, and F1-score. Additionally, we fine-tuned BioClinicalBERT using VA and I2B2 training data to assess the transferability of transformer models across public and private datasets.

**Results:** Transformer-based models outperformed rule-based tools when identifying names and locations, while rule-based systems demonstrated higher recall for more structured entities such as dates. CliniDeID showed the highest performance among external tools, but no tool achieved optimal balance across all PHI categories. Transformer models trained on both public and VA datasets performed best, though challenges remained in handling VA-specific terminology and annotations.

**Discussion:** These findings underscore the limitations of applying publicly trained models to private healthcare systems without adaptation. Differences in text structure, clinical terminology, and annotation standards impact deidentification effectiveness. While transformer models offer superior flexibility, computational cost and privacy concerns may hinder large-scale adoption.

**Conclusion:** The results emphasize the need for institution-specific evaluation and fine-tuning of deidentification models. Future work should explore large language model and hybrid approaches that integrate rule-based and transformer-based methods to enhance adaptability and accuracy in diverse clinical settings.

## INTRODUCTION

Clinical notes contain valuable clinical observations for research and patient care that is often missing or incomplete elsewhere in an electronic health record (EHR). However, their unstructured nature also introduces concerns related to patient privacy. Natural language processing (NLP) is able to extract and map information from clinical notes to specific clinical variables to allow integrated use with structured data. NLP methods have been used successfully in a clinical setting for research and operations purposes for decades[1, 2, 3]. With increased adoption of NLP methods into EHR research, protection of patient data located in text remains paramount. While prior studies have explored various approaches to clinical note deidentification, a better understanding of how these approaches and more contemporary approaches transfer between public and private data sources is needed.

The Health Insurance Portability and Accountability Act of 1996 (HIPAA) established standards in the United States for use and protection of patient data in the EHR. HIPAA’s Privacy Rule outlines definition of protected health information (PHI) as individually identifiable health information of a covered entity such as a patient[4] and deidentification of PHI as health data for which there is no reasonable basis to believe it can individually identify the subject[5]. The process of deidentification itself is defined either as expert evaluation and removal of identifiable information, or the “Safe Harbor” method as the removal of eighteen identifiers such as names, dates, and social security numbers referring to the subject and their relatives, employers, or household members from health data[5].

The variance in expression of concepts in text makes accurate removal of PHI difficult. NLP methods for deidentification have evolved over the years, but the scope of evaluation is limited by the same data protection rules being studied. Deidentification performance is typically measured on a single healthcare organization or a publicly available data set, such as those from Informatics for Integrating Biology and the Bedside (I2B2) [6, 7], with more recent work incorporating multiple sites and addressing these complexities [8]The difficulty of accessing clinical notes beyond public datasets and one or two healthcare organizations means most clinical note deidentification solutions are only evaluated using notes at the same organizations in which they were developed.

The US Department of Veterans Affairs (VA) is a health care system with 1,961 locations in the United States, US territories, and the Philippines. Thirty years of EHR data for over 25 million patients is stored in the VA Corporate Data Warehouse (CDW) for downstream use. As one of the largest integrated health care systems in the country, the data from VA patients enable many EHR-based studies. Some significant VA studies, such as those affiliated with the Million Veteran Program (MVP),[9] protect patient identities by providing VA-affiliated researchers with deidentified clinical data, in which clinical notes are rarely included. Prior studies evaluated and developed deidentification tools for VA clinical notes[10, 11]; however, the most common NLP methods and the scope of deidentification needs at VA have changed substantially since these evaluations have been performed.

The increased use of VA clinical notes for research purposes and the continued success of MVP research projects have emphasized the need for note deidentification and patient data protection at VA. In this paper, we describe an annotated corpus of VA clinical notes to compare and evaluate several existing deidentification methods and provide considerations when using deidentification tools on VA data. We additionally provide lessons learned for evaluating transformer models using combinations of public and private annotated data sources.

## METHODS

### Document Selection and Annotation

We sampled 1,000 clinical notes from the VA CDW, which contains notes from all clinical specialties and a variety of other VA services such as social work. Notes were randomly sampled from a large subset of VA notes evenly distributed among VA hospitals and years. The text at VA is diverse from many different specialties and services, and many notes do not have any PHI. To increase the number of notes sampled that likely contained PHI, we excluded nursing notes, addenda to other notes, and temporary notes. The resulting note types and clinical specialties can be seen in Table 1. Document metadata, including the document title, stop codes (VA billing codes), and author specialty, were used to categorize the notes. The highest proportion of notes were from the primary care domain (37.8%). We split these 1,000 notes into a 550 note training set, 50 note validation set, and 400 note held-out test set.

**Table 1:** Distribution of note types for the 1,000 VA notes sampled.

| **Note Category** | **Count** |
| --- | --- |
| **Administrative** | 108 |
| **Diagnostic/Ancillary Care** | 130 |
| **Emergency Medicine** | 46 |
| **Mental Health** | 61 |
| **Other Inpatient** | 6 |
| **Other Outpatient** | 35 |
| **Other Specialty Care** | 99 |
| **Primary Care** | 378 |
| **Rehabilitation Care** | 80 |
| **Missing/Unknown** | 57 |

Expert clinical annotators annotated the notes using the eHOST annotation tool[12]. The annotation guideline was based off the guideline used to create the I2B2/UTHealth 2014 shared task on clinical note deidentification[7]. Each instance of PHI was labeled with one of seven types: Name, Profession, Location, Age, Date, Contact, or ID. These PHI types cover more information than what is required by the HIPAA Safe Harbor method.

Clinical annotators went through multiple training rounds annotating notes to ensure annotation quality and agreement before annotating the final corpus. In the final note sample, annotators double annotated 100 notes from the corpus to evaluate inter-annotator agreement and ensure quality.

### External Tool Evaluation

We evaluated our annotated notes against four off-the-shelf tools for clinical note deidentification. Tools were selected based on three requirements: the tool had to be open source, it had work offline, and it had to output the identified PHI for evaluation, rather than only transforming the clinical text.

Based on these requirements, we selected Physionet Deid[13], Philter[14], CliniDeID[15], and Stanford Deid[16]. Physionet Deid and Philter both use rule-based methods; Physionet Deid relies on curated lists of known PHI terms and patterns, and Philter augments lists of PHI terms with lists of known “safe” or non-PHI terms and labels unknown words as PHI. CliniDeID is a hybrid system, combining term lists with simple machine learning based models. Stanford Deid is a transformer-based model. Due to not meeting one or more of the stated requirements, Amazon Medical Comprehend[17], John Snow Labs SparkNLP for Healthcare[18], CLAMP[19], and the National Library of Medicine’s Scrubber[20] were not considered at this time. Each tool processed the 400 notes in our heldout evaluation set using default settings (Physionet Deid, Philter, Stanford Deid) or settings to maximally capture PHI (CliniDeID) based on which would most closely capture the PHI described in the I2B2 annotation guideline. The tools did not label some of the output PHI types in the same way as our annotation guideline, so tool output was mapped to equivalent types in our annotated dataset where appropriate. In order to accurately compare tool performance, output types with no equivalent annotation type were excluded from evaluation.

### Fine-Tuned Transformer Models

Given the rapid change in NLP methods and state-of-the-art performance of transformer-based models in named entity recognition (NER), we also conducted several experiments utilizing a subset of these notes to identify PHI by fine-tuning BioClinicalBERT[21].

Four models were trained using different training methods on the two training sets:

1. **VA**: Trained on 550 VA training notes, evaluated with early stopping on 50 VA validation notes.
2. **I2B2**: Trained on the 792 I2B2 2014 training notes, early stopping on the I2B2 validation set.
3. **I2B2+VA**: Trained using the combined VA and I2B2 training sets, evaluated with early stopping using the VA and I2B2 validation notes combined.
4. **I2B2+VA FT**: Fine tuned the **I2B2** further using the VA training set and evaluating early stopping with the VA validation set.

All models employed token classification with a Conditional Random Field (CRF) head for prediction. Input and predicted output for each entity utilized BIO (Begin, Inside, Outside) format[22]. Model weights were initialized using BioClinicalBERT, with a maximum length of 128 tokens, and a learning rate of 1e-5.

## RESULTS

### Annotations

Of the 1,000 total notes, 197 (19.7%) had no PHI identified by the annotators. There were 7,413 instances of PHI annotated in the remaining 803 notes, with 4,765 (64.3%) of these annotations representing dates. Excluding dates, names and locations make up 1,870 (70.6%) of the remaining 2,648 annotations. Distribution of PHI types can be seen in Table 2.

**Table 2:**
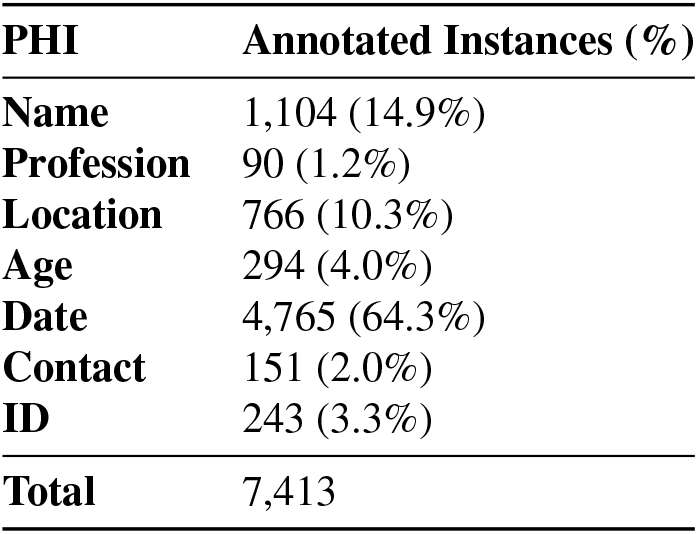
Annotation results of 1000 VA clinical notes based on the I2B2 2014 guideline.

Of the 100 documents that were double annotated, agreement between the two clinical annotators was 0.9526. Areas of annotator disagreement primarily included dates, ages, and professions. Notes with annotator disagreement were much longer than those with no disagreement, with an average length of 3,664.32 characters in notes containing disagreement versus an average length of 1,744.28 characters in the overall corpus.

### External Tool Evaluations

In table 3 we can see that CliniDeID was the best performing external tool on VA test set, with an F1 of 0.8422. Philter, Physionet Deid, and CliniDeID all have high recall; however, the Stanford Deid tool’s recall suffers due to a low performance on dates in our test corpus, with a date-specific recall of 0.5935.

**Table 3:** Performance of open source deidentification tools and fine-tuned Bio-ClinicalBERT models with results calculated using all PHI types and all PHI types except dates.

|  | Including Dates |  |  | Excluding Dates |  |  |
| --- | --- | --- | --- | --- | --- | --- |
|  | Precision | Recall | F1 | Precision | Recall | F1 |
| **Physionet Deid** | 0.6804 | 0.8074 | 0.7385 | 0.4816 | 0.6884 | 0.5668 |
| **Philter** | 0.4024 | 0.8467 | 0.5456 | 0.2056 | **0.7584** | 0.3235 |
| **CliniDeID** | 0.7529 | **0.9555** | **0.8422** | 0.5074 | 0.7439 | 0.6033 |
| **Stanford Deid** | **0.8205** | 0.6105 | 0.7001 | **0.6499** | 0.6415 | **0.6457** |
| **VA** | 0.8870 | 0.8650 | 0.8758 | 0.7824 | 0.7731 | 0.7777 |
| **I2B2** | 0.7616 | **0.9076** | 0.8282 | 0.5450 | **0.8503** | 0.6642 |
| **I2B2+VA** | 0.8796 | 0.7861 | 0.8302 | 0.5891 | 0.7467 | 0.6586 |
| **I2B2+VA FT** | **0.9120** | 0.8942 | **0.9030** | **0.8196** | 0.8132 | **0.8164** |

Since dates make up 64.3% of our annotated data, a tool’s performance on dates alone could obfuscate performance on the other PHI types. Therefore, we additionally calculated performance for the tools while excluding date types from evaluation.

All evaluated tools benefit substantially from a large portion of annotated PHI representing dates. CliniDeID had the largest reduction in overall performance, dropping from an F1 of 0.8422 to 0.6033, a decrease of 28.3%, but Philter remained the lowest performing external tool, with F1 dropping from 0.5456 to 0.3435. Stanford Deid had the lowest performance drop, largely due to its already low performance on dates already impacting its F1. After removing dates, Stanford Deid’s performance is the highest in both precision and F1, indicating that the pre-trained transformer model is more robust to complex VA data than the rule-based tools, although its recall is the lowest of all tools evaluated.

### Fine-Tuned Transformer Models

The fine-tuned BioClinicalBERT models performed comparably or better than all evaluated externally developed tools. The **I2B2+VA FT** model showed the highest performance in precision and F1 when evaluating with and without dates. However, the **I2B2** model had the highest recall in both evaluations.

The fine-tuned BioClinicalBERT models suffer performance losses when excluding dates from evaluation, but not as significantly as the external tools. The **I2B2+VA FT** model has the lowest performance drop, going from an F1 of 0.9030 to 0.8164, an 9.6% drop. The performance for **VA** is balanced evenly between both precision and recall for both evaluations.

## DISCUSSION

This study evaluated several clinical note deidentification tools and transformer-based models. These four tools use a variety of deidentification methods. Two of the selected tools, Physionet Deid and Philter, are rule-based. Physionet Deid relies exclusively on curated lists of known PHI terms and patterns. Philter augments lists of PHI terms with lists of known “safe” or non-PHI terms and labels unknown words as PHI. CliniDeID is a hybrid system, combining term lists with simple machine learning based models. Stanford Deid is a transformer-based model similar to our BioClinicalBERT experiments. Our four fine-tuned BioClinicalBERT models show the transferability of utilizing existing public datasets as a training source and the impact of augmenting existing datasets with VA data.

CliniDeID and the four transformer-based models performed best under most of our evaluations. Philter and Physionet Deid, the two rule-based solutions, had the least accurate predictions when applied to VA clinical notes. Philter in particular was prone to over-prediction, with eight times as many predicted names as were in the annotated test data, largely due to its labelling of unknown terms as PHI. This resulted in Philter having recall equivalent to the other tools at the expense of both its precision and F1 measures being at least 26% lower than all other methods evaluated.

The experiments using the **VA** BioClinicalBERT models’ reveal that the transformer based models have a high transferability between different data sources, with only minor differences in F1 scores. This suggests that in cases where there is no annotated data available for a private data source, the models fine tuned on public data source can still be applied with relatively better performance than the rule based models in this task.

Additionally, the model fine-tuned on public data and then further trained on private data outperformed a model trained on a mixed public-private dataset. This difference may be due to the use of different validation datasets, with the former using only the private validation dataset, and the latter using both public and private data within the validation dataset. The combined validation dataset could lead the trained model toward a more generalized model, while the continue trained model only uses private data resulting the model performing better on testing only private data. However, more experimentation is required to further investigate the factors contributing to observed performance between these models.

All of these evaluated tools come with a variety of practical challenges that need to be overcome for widespread use for VA research. The size and diversity of the VA corpus means broad adaptation of rule-based vocabularies to VA notes is likely infeasible and targeted improvement of the most frequent or critical error types would be necessary. Notably, all evaluated systems show large improvements in the deidentification task over the last decade since similar work was performed at the VA[10]. Although our experiments fine-tuning BioClinicalBERT showed good performance and all four transformer-based models had better performance on non-date PHI types than other methods, practical implementation of these models at scale for VA research remains untested due to compute availability and cost.

The annotated corpus in this work as well as in similar corpora such as in I2B2 2006[6], I2B2 2014[7], and in prior evaluation of deidentification tools at VA[10], all reveal that dates are the most prevalent form of annoted PHI. However, dates also have limited textual and are well-captured by the evaluated tools and models with the exception of Stanford Deid. Removing dates from the evaluation process places a greater emphasis on names and locations, which are more variable annotated data types.

The all-veteran population at VA healthcare centers introduces a variety of extra considerations to deidentification. One problem that appeared in our experiments was that the word “veteran” is used interchangeably with “patient” and can occur dozens of times in a single note. CliniDeID detects all instances of the word “veteran” as an organization or profession PHI type. While this may be appropriate in other contexts, this is not present in our annotated data.

Additionally, HIPAA’s Safe Harbor deidentification requirements include “any other unique identifying number, characteristic, or code” covering other forms of identifiable information not otherwise listed. At VA, a patient’s military branch, rank, or details about dates or locations of deployment are sometimes found in notes and may be identifiable information. Even if this information is removed, other secondary information may infer the removed data. A patient with Agent Orange exposure documented in a deidentified note is identifiable to the Vietnam War, regardless of whether explicit mentions of deployment in Vietnam have been removed. Although these types of information are not always individually identifying, they would need to be considered for any public or semi-public release of VA clinical notes such as for an I2B2 shared task or any other public annotated data.

Internal VA use of deidentification may not require as much data deidentified as public release of data. Our annotated data and all evaluated NLP tools remove more data than is required by HIPAA. MVP removes birth dates, but includes dates other dates such as any diagnosis, admission, discharge, and military service. In this paper, we did not distinguish any date in the text from date of birth of a patient. Although CliniDeID disambiguates dates of birth from other dates, the accuracy of this disambiguation is not measured.

Due to some variance in the development goals and annotation guidelines, re-evaluating based only on PHI from the HIPAA “Safe Harbor” requirements may improve performance on certain under-predicted categories with more narrow HIPAA requirements like locations and ages. Configuration of each tool is also an avenue that was not explored at this time. Most tools have some customization available at runtime, with CliniDeID having sophisticated configuration of all PHI types that may impact performance.

## CONCLUSION

All the evaluated methods for clinical note deidentification have significant drawbacks when applied to VA notes that underscore the importance of evaluation when applying externally developed tools to site specific data. A relatively small test set, smaller than the I2B2 2014 test set, was sufficient to identify weaknesses of external tools on VA data. This work establishes a foundation for future benchmarking of additional LLM-based deidentification approaches, which may offer further improvements but require additional validation and considerations within the constraints in the VA setting.

All externally developed tools suffer performance losses when evaluating more complex PHI types such as names and locations, with the four fine-tuned Bio-ClinicalBERT models performing the best on these complex PHI. Although the high overall performance of the transformer-based models is promising, issues with compute availability and cost will slow their adoption for deidentification of large corpora for VA research. Some errors may be addressed with post processing or utilizing a tool’s built-in configurations.

Even when using the HIPAA Safe Harbor requirements as a guide, determining what is and is not PHI remains complicated. Additional differences, such as institutional requirements or study-specific deidentification decisions, impede the cross-compatibility of any developed dataset or solution. VA-specific identifying information is not well-handled by any externally developed solution or annotation guideline.

## Data Availability

The data for this study originates from the U.S. Department of Veterans Affairs and includes protected health information (PHI); therefore, it is not available for public distribution.

## ACKNOWLEDGEMENTS

This work was supported using resources and facilities of the Department of Veterans Affairs (VA) Informatics and Computing Infrastructure (VINCI), funded under the research priority to Put VA Data to Work for Veterans (VA ORD 22-D4V). The views expressed are those of the authors and do not necessarily represent the views or policy of the Department of Veterans Affairs or the United States Government.

